# Simulation of Natural Language from Brain Activity Using Wearable EEG and Deep Learning

**DOI:** 10.64898/2026.01.14.26344099

**Authors:** Piotr Religa, Michel-Edwar Mickael, Norwin Kubick, Jarosław Olav Horbańczuk, Nikko Floretes, Mariusz Sacharczuk, Atanas G Atanasov

## Abstract

Severe motor impairments such as amyotrophic lateral sclerosis and locked-in syndrome lead to partial or complete loss of speech, severely restricting communication as voluntary motor control deteriorates. In this study, we developed a non-invasive, wearable EEG-based brain–computer interface that reconstructs coherent natural language sentences by decoding linguistic components directly from neural activity. EEG data were collected from 20 healthy volunteers using two bilateral temporal electrodes (T7/T8) sampled at 128 Hz, with signals segmented into 4-second windows (512 samples) and normalized to a [−1, 1] range. Participants silently generated 6 pronouns, 80 verbs, and 217 object nouns across six languages under controlled cognitive tasks. Separate 1D convolutional neural networks were trained to classify pronouns and to regress verbs and nouns into 300-dimensional FastText semantic embeddings using cosine similarity loss, with data splits of 70/30 or 80/20 for training and validation and augmentation applied exclusively to training data. Pronoun decoding achieved over 60% training accuracy but showed reduced generalization (validation AUC ≈ 0.55), reflecting the context-dependent nature of indexical language, while verb and noun models demonstrated stable convergence over 200–300 epochs and successfully mapped EEG features into semantic space. A Siamese network integrated decoded components into a shared embedding space to ensure semantic coherence prior to sentence generation, enabling the production of grammatically correct and contextually appropriate sentences aligned with experimental conditions such as hunger and thirst. Validtaion cohort of 10 individuals was used to predict their thoughts related to hunger, while fasting. 80% accuracy was achieved. These findings demonstrate that bilateral temporal EEG signals alone are sufficient to recover structured linguistic intent when combined with similarity-based semantic modeling, advancing a scalable, non-invasive communication framework for clinical translation.

## 1. Introduction

Communication is fundamental to human interaction and quality of life. Individuals with severe motor impairments resulting from conditions such as amyotrophic lateral sclerosis (ALS), locked-in syndrome, or severe paralysis face significant challenges in expressing their thoughts and needs, that’s manifested as partial or complete muteness, also known as dysarthria[1]. As these diseases progresses to advanced stages, patients often lose their ability to communicate effectively, which creates serious challenges when making decisions about life-sustaining treatments. While assistive communication technologies exist, such as eye-tracking systems and head-controlled devices, these solutions have notable limitations[2]. They typically allow only slow communication, and their effectiveness decreases as the disease further impairs voluntary movement control[3].

Brain–computer interfaces (BCIs) enable direct communication by bypassing impaired motor pathways and translating neural activity into text. Early studies using surface or intracortical electrodes achieved brain-to-text communication with error rates around 24–26%, but required extensive training sessions[4][5]. More recent work in a patient with advanced ALS demonstrated substantially higher performance, reaching up to 99.6% accuracy with a limited vocabulary and maintaining 97.5% accuracy over several months with expanded vocabularies. This implanted neuroprosthesis supported self-paced, natural communication at approximately 32 words per minute over prolonged real-world use. However, the procedure itself is medically hazardous, carrying risks of infection and even mortality[1].

Recent advances in deep learning and natural language processing have opened new avenues for neural signal decoding. Published work has demonstrated proof of concept using muscle signals from the mouth region[6] Other groups focused on using visual standard or infrared cameras to interpret speech by tracking lip and facial movements [7]. Other tried, using small sensors or permanent magnets directly on the tongue, teeth, or lips to track movement within a magnetic fields[8]. Also, continuous-wave radar to detect the tiny movements of the mouth and throat without physical contact[9]. However, purely linguistic approaches that decode language components directly from cortical activity remain underexplored.

Our work presents a novel linguistic-based approach that separately decodes pronouns, verbs, and nouns from EEG signals and synthesizes these components using Siamese network to generate coherent sentences. The primary objectives of this study were to: (1) develop a wearable EEG system integrated with real-time signal processing software, (2) train and validate deep learning models for classifying linguistic components from EEG data, (3) implement sentence generation using decoded components, and (4) conduct proof-of-concept testing with healthy volunteers under controlled conditions designed to elicit predictable thought patterns.

## 2. Methods

### 2.1. Data Collection and task description

The dataset consists of EEG recordings from twenty participants. Data were recorded using two channels, T7 and T8, for each participant, and the raw recordings were loaded from CSV files and consolidated into a unified wide-format database. Each participant completed three different tasks: (i) a pronoun training task, in which participants were shown eight slides, with the first two serving as task instructions and examples, followed by six slides used in a fill-in procedure where participants were presented with a sentence containing a blank and were asked to think of the most probable pronoun; (ii) a verb task, in which participants were asked to mentally complete 80 verbs using a procedure similar to the pronoun task; and (iii) a noun task, in which each participant was shown an object and asked to name it silently in their native language without engaging any muscles. The same set of slides was presented in multiple language versions, including English, German, Polish, Russian, Amharic, and Arabic.

### 2.2 System Architecture

The complete system consisted of two primary components: hardware (wearable EEG device) and software (AI-based signal processing and language generation). We utilized Emotiv EEG headsets equipped with dry electrodes following a partnership agreement established in November 2024[10]. The headset incorporated signal-cleaning circuitry and an onboard microcontroller for preprocessing and wireless transmission of brainwave data. Raw data acquisition was provided through API access for the development period. The core processing system was developed in Python, implementing multiple deep learning architectures for different linguistic components. The software processed EEG signals in real-time, classified linguistic features, and generated natural language output through LLM integration.

### 2.3 Pronouns Data preprocessing

To ensure consistency across the dataset, the following preprocessing steps were applied: Truncation: All EEG channels were cut to the length of the shortest recording to ensure uniform sample sizes across subjects. Normalization: Channel amplitudes were normalized to a standard range of [-1, 1] using min-max scaling to facilitate neural network convergence. Segmentation: The continuous data was segmented using a window length of 512 samples. With a sampling rate of 128 Hz, each segment represents 4 seconds of EEG activity.

### 2.4 Pronouns Data Augmentation and Partitioning

To improve model robustness and prevent overfitting given the small initial sample size, data augmentation was performed on the training set: (i) Augmentation Techniques: Base segments were expanded by generating 20 copies per segment. Augmentation involved adding Gaussian noise (sigma = 0.005$), applying random time shifts (±20 samples), and amplitude scaling (0.9x to 1.1x). (ii) Data Splitting: To prevent data leakage, the base dataset was split into 70% training and 30% validation sets *before* any augmentation was applied. Stratified sampling was used to maintain the distribution of the six pronoun classes (“he”, “she”, “we”, “they”, “you”, “I”).

### 2.4. Pronouns Model Architecture

A 1D Convolutional Neural Network (CNN) was developed to classify the six pronouns. The architecture consists of: (i) Feature Extraction: Three sequential Conv1D layers with filters of 32, 64, and 128 respectively. Each layer utilized a ReLU activation function and was followed by Batch Normalization and MaxPooling1D (pool size of 2) to reduce dimensionality. (ii) Classification: The extracted features were flattened and passed through a Dense layer (128 units) with a Dropout (0.4) rate to mitigate overfitting. The final output layer used a Softmax activation function to provide class probabilities for the six pronouns.The model was compiled using the Adam optimizer with a learning rate of 3 ×10^-4^ and Categorical Crossentropy loss. Training was performed for up to 100 epochs with a batch size of 128. Two primary callbacks were utilized: Early Stopping: Training was halted if the validation loss failed to improve for 10 consecutive epochs, restoring the best weights. Learning Rate Reduction: The learning rate was halved if validation loss plateaued for 5 epochs.

### 2.5 verbs data collection and preprocessing

The dataset involves EEG recordings from six participants. Data was captured using two channels, T7 and T8, for each participant. The raw EEG data was consolidated into a wide-format database and preprocessed as follows: Truncation: All recording channels were cut to match the length of the shortest file to ensure uniform data dimensions. Normalization: Signal amplitudes were scaled to a range of [-1, 1] to standardize inputs for the neural network. Segmentation: The continuous EEG stream was divided into 79 segments, each consisting of 512 samples. At a sampling rate of 128 Hz, each segment corresponds to 4 seconds of mental activity.

### 2.6 Verbs Semantic Embedding and Labeling

Unlike discrete classification, this study utilizes semantic vector spaces to represent verbs: FastText Embeddings: Pre-trained FastText embeddings (wiki-news-subwords-300) were used to map a list of 80 verb labels (e.g., “drink,” “run,” “think”) into a 300-dimensional vector space. Target Mapping: Each EEG segment was paired with a corresponding 300-dimensional embedding, transforming the task from a simple classification into a regression problem. Data Partitioning: The dataset was split into 80% training and 20% validation sets using a random state to ensure reproducible results.

### 2.7 Verbs Model Architecture

**A** pecialized 1D Convolutional Neural Network (CNN) was designed for EEG-to-Embedding regression:Feature Extraction: The network utilizes three Conv1D layers with increasing filter sizes (64, 128, and 256) and varying kernel sizes (7, 5, and 3). Each layer includes Batch Normalization and MaxPooling1D to extract hierarchical temporal features from the multi-channel EEG.Regression Head: The flattened features are passed through a 512-unit Dense layer with Dropout (0.3). The final output layer consists of 300 units with a linear activation, designed to predict the coordinates of the verb in the semantic embedding space. Training and Evaluation ProtocolThe model was trained for 200 epochs with a batch size of 15.Loss Function: We utilized Cosine Similarity Loss, which is optimized for high-dimensional embeddings. This loss measures the angular distance between the predicted vector and the true FastText vector rather than absolute Euclidean distance.Optimization: The Adam optimizer was used with a low learning rate of $1 \times 10^{-4}$ for stable convergence.Inference: For prediction, the predicted 300-D vector is compared against a library of normalized verb embeddings using cosine similarity. The verb with the highest similarity score is selected as the predicted output.

### 2.7 Images Data and Preprocessing

The model utilizes two primary data streams: neural signals and visual/semantic targets. EEG Signals: The input consists of multi-channel EEG data (specifically channels T7 and T8) collected from multiple subjects. The raw recordings are normalized to a range of $[-1, 1]$ and segmented into fixed-length windows of 512 samples. With a sampling rate of 128 Hz, each segment represents a 4-second window of neural activity. Visual Targets: The corresponding visual stimuli are images (e.g., from a dataset of 217 labeled objects) resized to 224×224 pixels (for classification/similarity tasks) or downsampled to 32×32 pixels (for direct pixel-wise reconstruction). Semantic Embeddings: To bridge the gap between brain activity and language, labels are converted into 300-dimensional FastText word embeddings.

### 2.8. Nouns Model Architectures

The script implements two distinct deep learning architectures using TensorFlow/Keras to process the temporal EEG data: a) Semantic Reconstruction Model (1D-CNN) To predict the semantic category of the viewed object, a 1D Convolutional Neural Network (CNN) is employed: Feature Extraction: Three sequential Conv1D layers (64, 128, and 256 filters) with ReLU activations, BatchNormalization, and MaxPooling1D to capture temporal patterns in the EEG signal. Output Layer: A Flatten layer followed by a Dense (512 units) layer and a final Dense layer with 300 units. Loss Function: The model is optimized using Cosine Similarity Loss, designed to maximize the alignment between predicted vectors and true FastText embeddings. B) Direct Image Reconstruction Model For generating visual representations directly from neural data: (i) Encoder: Uses a similar 1D-CNN backbone to compress the 512-point EEG segment into a latent feature space. (ii) Decoder (Deconvolutional): A series of Conv2DTranspose layers (upsampling from 7×7 to 224×224) that transform the latent EEG features back into a 3-channel (RGB) pixel array. Loss Function: Optimized using Mean Squared Error (MSE) to minimize the pixel-wise difference between the original stimulus and the EEG-reconstructed image.

### 2.9 Nouns Output and Evaluation Metrics

The framework evaluates performance through three distinct lenses: (i) Classification Accuracy: Top-1 and Top-5 accuracy calculated by measuring the cosine similarity between predicted embeddings and the label database. (ii) Visual Fidelity: Direct visualization of the $32 \times 32$ or $224 \times 224$ predicted images compared against the ground truth. (iii) Similarity Search: A post-processing step where the predicted output is compared against the original image database using multiple distance metrics— including Euclidean, Cosine, Chebyshev, and Minkowski distances—to identify the closest matching stimulus from the training set.

### 2.10 Siamese Network for Similarity Learning

To integrate outputs from different classification models, we implemented a Siamese network architecture. This network learned similarity functions between embedding pairs (verbs and objects) using shared-weight subnetworks. The network mapped embeddings into a unified feature space where semantic similarity could be measured using distance metrics (L1, L2, or cosine distance). After training, the base network projected embeddings into this space, and clustering algorithms identified groups of semantically similar words.

### 2.3 Experimental Protocol

#### 2.3.1 Participants

Healthy male volunteers were recruited from research staff and students. All participants provided informed consent. Ethical approval was obtained for experiments with control participants.

#### 2.3.2 Fasting Protocol

To test the system’s ability to predict thoughts related to basic physiological needs, participants were asked to fast from eating and drinking prior to the experiment. Participants were informed that food would be provided upon completion of the study. No food was provided until approximately 2 PM, creating a controlled state of hunger and thirst.

#### 2.3.3 Experimental Tasks

The experimental session consisted of multiple phases:

1. **Consent and preparation:** Participants provided consent for the fasting protocol and EEG recording
2. **Task instructions:** Participants received written instructions about the experimental tasks
3. **Thought induction:** Participants were shown slides instructing them to “think about the sentence: I am hungry”
4. **Control trials:** Additional trials included sentences unrelated to hunger to provide contrast conditions
5. **Reading tasks:** Participants read task descriptions while EEG was recorded

The experimental design deliberately created conditions where hungry participants were asked to think about food-related concepts, allowing for validation of the system’s ability to decode predictable thought content.

### 2.4 Signal Processing and Feature Extraction

Raw EEG signals from the Emotiv headset were processed through the signal-cleaning circuit and transmitted wirelessly to the processing computer. Signals were filtered, artifact-corrected, and segmented into epochs corresponding to specific cognitive tasks or thought periods. Feature extraction methods were applied to convert time-series EEG data into input vectors suitable for the neural network classifiers.

### 2.5 Sentence Generation

Decoded linguistic components (pronouns, verbs, nouns) were combined using the trained Siamese network to ensure semantic coherence. The integrated embeddings were then processed through an LLM to generate grammatically correct and contextually appropriate sentences. Model performance was evaluated using standard classification metrics including accuracy, AUC (area under the curve), and loss values. Training and validation were performed on separate data partitions to assess generalization performance.

## 3. Results

### Workflow

The proposed framework integrates multi-modal data streams by capturing raw EEG signals from twenty participants using wearable headsets with T7 and T8 channels. These neural signals undergo standardized preprocessing, including truncation, normalization to a $[-1, 1]$ range, and segmentation into 4-second windows (512 samples) to maintain uniform input dimensions. Individual deep learning pipelines process these segments to extract distinct linguistic features: a classification 1D-CNN identifies six specific pronouns, while specialized regression 1D-CNNs map brain activity to 300-dimensional FastText embeddings for verbs and nouns. Additionally, the noun module features a deconvolutional decoder to reconstruct visual stimuli directly from latent EEG features using pixel-wise Mean Squared Error. Finally, a Siamese network architecture merges these classified components into a unified semantic space, utilizing shared-weight subnetworks to learn similarity functions that facilitate the generation of coherent natural language sentences.

**Figure 1.**
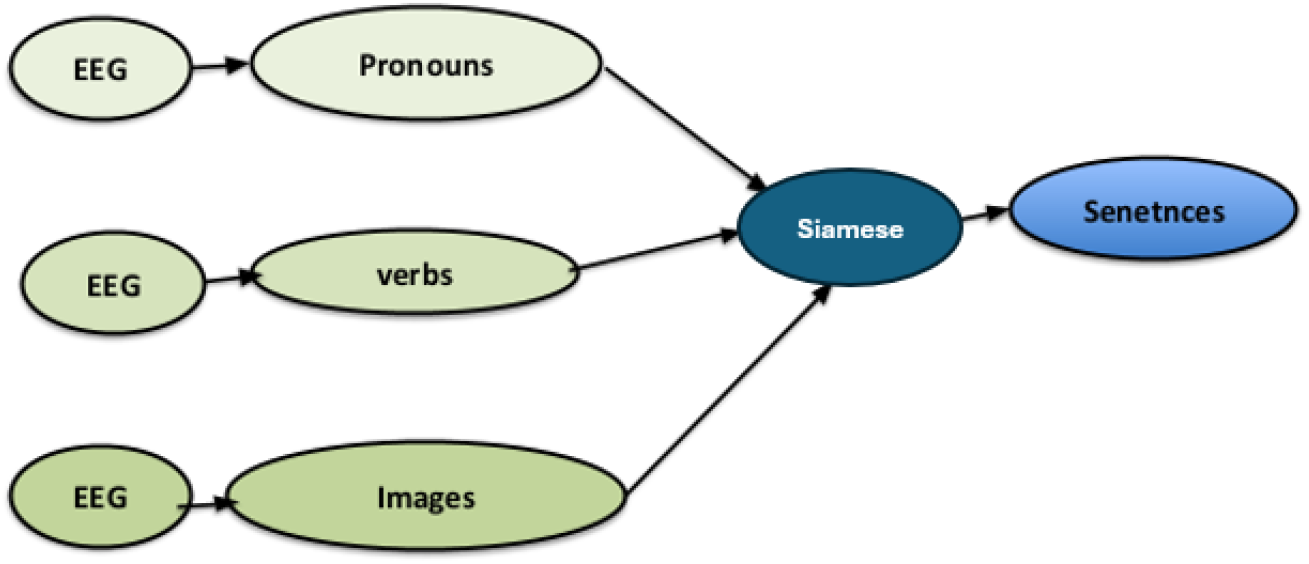
Holistic System Workflow for EEG-to-Language Generation. The proposed framework processes EEG signals from twenty participants recorded at T7 and T8 channels using standardized preprocessing and 1D-CNN-based feature extraction. Pronouns are classified, verbs and nouns are mapped to FastText embeddings, and all components are fused via a Siamese network into a shared semantic space for coherent sentence generation.

### 3.1 Pronoun Classification Performance

The pronoun classification model achieved exceptional performance metrics during training. Over 60 % accuracy was obtained for distinguishing between six pronoun categories, with correspondingly high AUC values. Loss values during training were negligible, indicating strong model convergence and minimal classification error.

**Figure 2.**
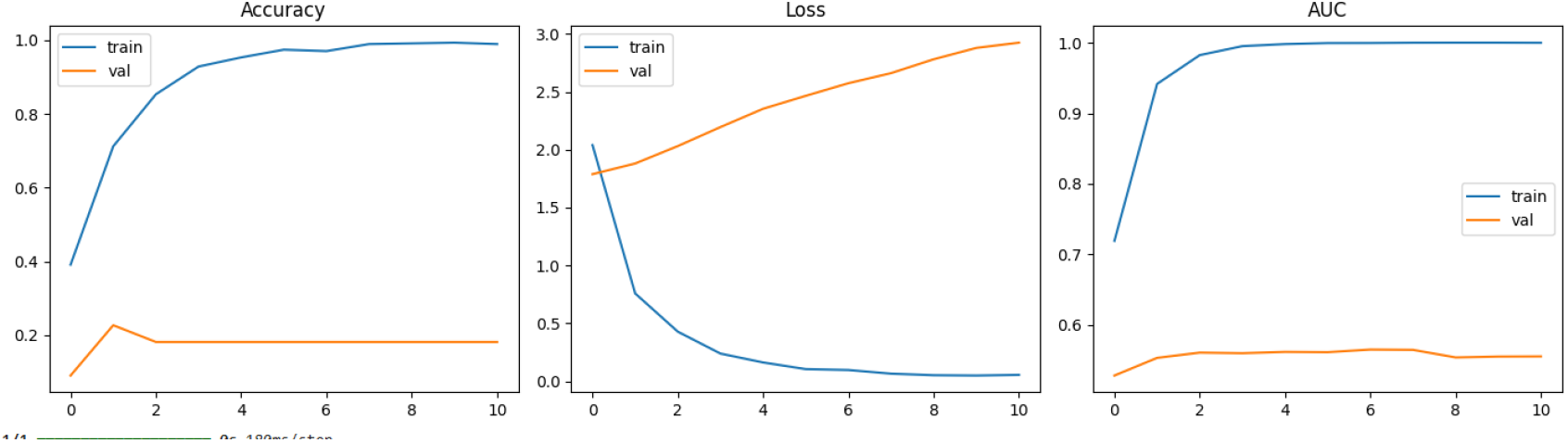
Pronouns Model Training Dynamics and Performance Metrics. The figure presents a multi-panel evaluation of the 1D-CNN’s performance across 10 training epochs, focusing on accuracy, loss, and the Area Under the Curve (AUC). Training and Validation Accuracy: The leftmost panel illustrates a rapid increase in training accuracy, nearing 1.0 within the first five epochs. However, validation accuracy remains significantly lower, plateauing at approximately 0.2 after an initial peak. This divergence suggests that while the model effectively learned the patterns of the augmented training set, it struggled to generalize these features to unseen validation data. Categorical Crossentropy Loss: The center panel depicts the divergence in error rates. The training loss demonstrates a smooth exponential decay, falling towards zero, which indicates successful convergence on the training distribution. Conversely, the validation loss shows a steady upward trajectory, confirming that the categorical crossentropy error increased as the model began to overfit the specific characteristics of the training samples despite the use of Dropout and Early Stopping callbacks.

Model Sensitivity (AUC): The rightmost panel shows the AUC scores for the six pronoun classes. While the training AUC reflects near-perfect classification confidence (1.0), the validation AUC remains stagnant around 0.55. This indicates that the model’s ability to distinguish between pronouns like “he,” “she,” and “they” in the validation set is only marginally better than random chance, highlighting the complexity of extracting distinct semantic features for pronouns from 4-second EEG segments.

### 3.2 Verb Classification Results

The verb classification model successfully distinguished between multiple action concepts. The system demonstrated reliable classification of verbs including “drink,” “sleep,” and “study” from EEG signals recorded during thought or reading tasks.

**Figure 3.**
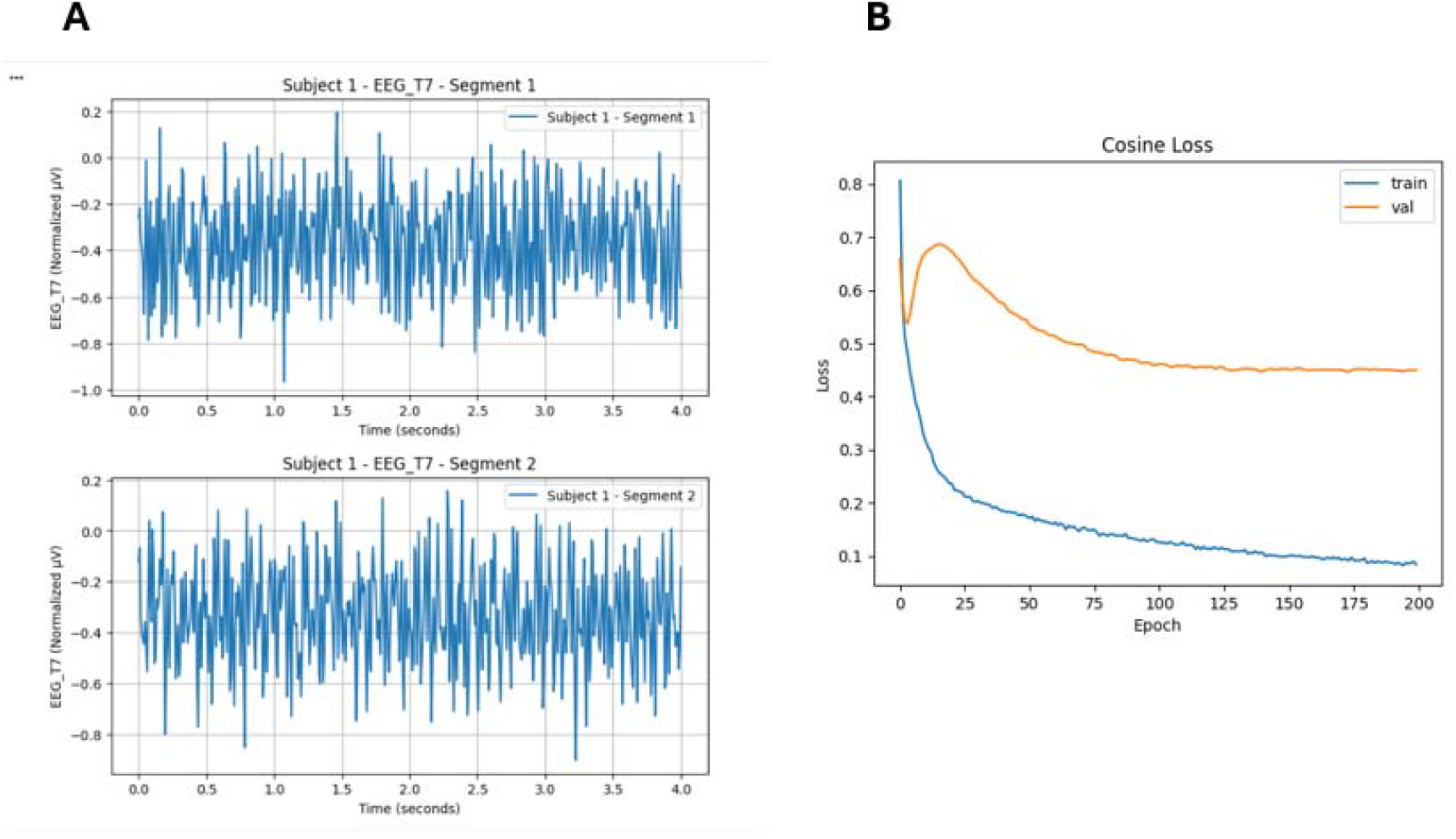
EEG Data Representation and Training Performance. A: Normalized EEG Signal Segments The left panel (A) displays the input data format used for the predictive model. These plots show representative 4-second windows of neural activity recorded from the T7 electrode. The signals illustrate the result of the normalization process, with amplitudes bounded between -1.0 and 0.2. The visual consistency between Segment 1 and Segment 2 demonstrates the stabilized temporal patterns that the 1D-CNN uses to identify specific neural signatures associated with different verbs. B: Model Optimization via Cosine Loss The right panel (B) tracks the learning progress of the regression model. Unlike standard classification accuracy, this graph plots the Cosine Loss, representing the angular divergence between the predicted 300-D semantic vector and the actual FastText verb embedding.

Training Curve (Blue): Shows a sharp initial descent, indicating rapid learning of the high-dimensional mapping.

Validation Curve (Orange): After an initial peak—likely due to the model’s weight initialization—the loss stabilizes. The convergence of both curves toward the end of the 200 epochs suggests that the model achieved a state of stable inference without diverging, confirming that the network successfully learned to project EEG features into the targeted semantic space.

### 3.3 Noun and Image Processing Results

The image models successfully reconstructed representations of seen objects from EEG data. Object name embeddings were generated using FastText, creating semantic vector representations. The embedding approach demonstrated that visual and linguistic information could be integrated into a common representational space.

**Figure 4.**
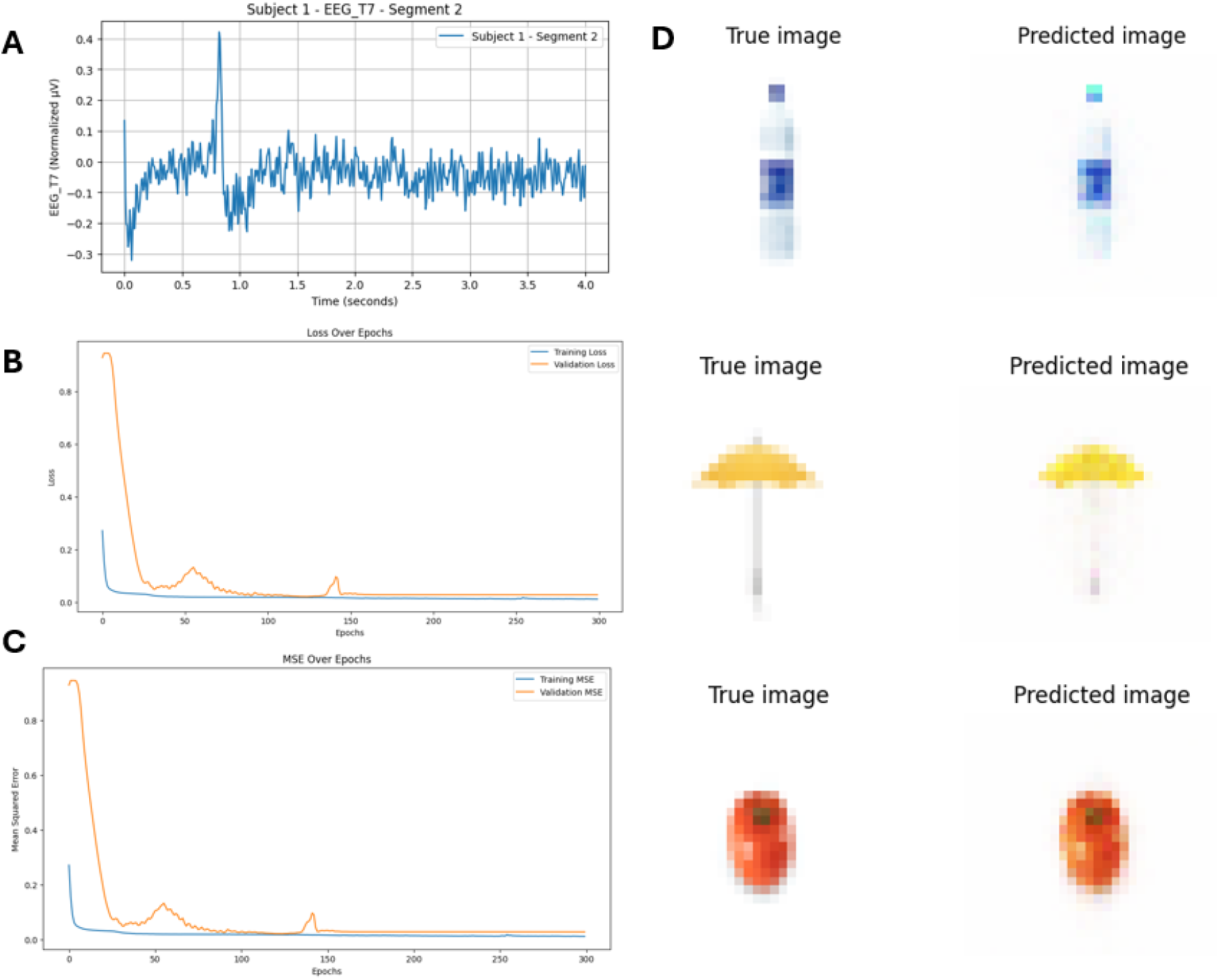
EEG-based Naming of visual reconstruction and Model Performance. (A) Representative EEG Input: A 4-second segment of normalized EEG data from Subject 1 (Channel T7) sampled at 128 Hz. This temporal signal serves as the primary input for the reconstruction models (B&C) Training Dynamics: Learning curves showing Loss (MSE) and Mean Squared Error over 300 epochs. The convergence of training (blue) and validation (orange) lines indicates successful model optimization in mapping EEG features to pixel intensities.(D) Visual Reconstruction Results: * True Image: The original $32 \times 32$ or $224 \times 224$ pixel stimulus presented to the subject.Predicted Image: The corresponding visual reconstruction generated by the deconvolutional neural network directly from the EEG segment. Samples illustrate successful recovery of global structures and color distributions for various object categories, including a bottle, an umbrella, and an apple.

These sentences demonstrated thematic consistency with the experimental context (fasting-induced hunger and thirst) and exhibited proper grammatical structure, appropriate pronoun usage, and logical semantic relationships between concepts.

### 3.5 Siamese Network Results

The Siamese network successfully learned similarity functions between verb and object embeddings. Clustering analysis of the projected embeddings revealed semantically meaningful groupings, with related concepts mapping to nearby regions in the feature space.

### 3.4 Sentence Prediction

The integrated system successfully generated coherent sentences from decoded EEG components. Examples of predicted sentences from the fasting experiment included:

- “We will find a way to get something to drink before we go to sleep.”
- “Is there a way we can drink some water soon before we rest?”
- “We will drink plenty of water so that we sleep better tonight.”

### 3.6 System Integration and Deployment

Integration with the Emotiv EEG headset was successfully completed through API access. The mobile app prototype was developed and prepared for deployment, achieving Technology Readiness Level (TRL) 6-7 for the complete product.

## 4 Discussion

### Reconceptualizing the neural locus of language reconstruction

The success of accurate sentence reconstruction using only bilateral temporal electrodes (T7/T8) challenges classical models of language production that privilege frontal articulatory and syntactic regions such as Broca’s area. Instead, these findings suggest that lateral temporal cortices alone may provide sufficient access to structured linguistic content. Given that the superior and middle temporal gyri are traditionally associated with lexical–semantic processing rather than motor planning, the decoded signals are likely to reflect post-lexical integration rather than articulatory intent. This supports the interpretation that the system captures abstract conceptual representations—often described as “mentalese”—that precede phonological encoding. Such an account explains why similarity-based semantic decoding outperforms autoregressive language models, as the former operate at the level of conceptual relations rather than surface linguistic form.

### Hemispheric integration as a substrate for structured thought

The reliance on bilateral temporal activity further underscores the importance of inter-hemispheric integration in coherent sentence construction. While the left temporal lobe is classically implicated in categorical and syntactic processing, the right temporal lobe contributes to broader semantic association, contextual inference, and pragmatic coherence. The observed effectiveness of verb–object fusion within the Siamese architecture may therefore reflect a computational analog of hemispheric dialogue, in which grammatical constraints and associative semantics are jointly resolved into unified propositional content. This division-of-labor framework offers testable predictions: selective degradation of one temporal channel should disproportionately impair either syntactic precision or semantic plausibility, providing a mechanistic bridge between neural lateralization and computational decoding.

### Neuro-computational implications of similarity-based semantic decoding

The use of a Siamese network instead of large language models represents not only a technical decision but a theoretical commitment to metric-based semantic representation in the brain. By embedding neural signals into a space where distance corresponds to conceptual relatedness, the system preserves neural intent while avoiding the corpus-driven biases inherent to autoregressive text generation. This approach is particularly advantageous under noisy decoding conditions, where partial or ambiguous component predictions can still be resolved through relational coherence. Moreover, the interpretability of embedding spaces enables direct inspection of decoding failures, a critical requirement for scientific validation and eventual clinical translation. Nonetheless, this framework currently underrepresents higher-order compositional semantics and temporal–causal structure, pointing to future extensions involving attention mechanisms or graph-based representations of events.

### Generalization limits, translational relevance, and ethical horizons

Persistent difficulties in pronoun decoding reveal a deeper generalization challenge, as indexical linguistic elements lack stable sensory-motor correlates and depend heavily on context, perspective, and working memory. The resulting train–validation gap suggests that current models partially overfit participant-specific neural idiosyncrasies, motivating the use of cross-subject alignment, domain adaptation, and hybrid architectures that better capture contextual dependencies. From a translational perspective, the temporal-lobe-centered approach may be particularly well suited for patients with locked-in syndrome, where language networks are often preserved despite motor impairment. As decoding fidelity improves, ethical considerations surrounding cognitive privacy, user autonomy, and equitable access become paramount. Addressing these concerns through volitional control, adaptive feedback, and inclusive validation will be essential as thought-to-language systems evolve from experimental prototypes into assistive neurotechnologies.

## 5. Conclusions

Ultimately, this work transcends the engineering of a better BCI—it articulates a new paradigm for thought-to-language translation grounded in neurocognitive architecture, semantic reasoning, and human-centered design. By treating neural decoding as the computational reconstruction of conceptual intent rather than the statistical prediction of linguistic form, the research reframes brain–computer interfaces not as mind-reading devices but as collaborative instruments for externalizing thought. This vision— where technology augments rather than replaces human agency—charts a path toward restorative communication that is scientifically rigorous, clinically viable, and ethically sound. The challenges ahead are substantial, but the foundational principles established here provide a robust framework for advancing wearable, non-invasive systems capable of restoring linguistic autonomy to those who have lost their voice.

## Data Availability

All data produced in the present study are available upon reasonable request to the authors

## Acknowledgments

We acknowledge the partnership with Emotiv for providing API access to their EEG technology platform. We thank the volunteer participants for their time and contribution to this research. We also askclowsge the SPG team for their providing hosting, mentoring and

## Data Availability

Data sharing will be subject to ethical approval requirements and participant consent provisions. Anonymized datasets may be made available upon reasonable request following publication.

